# Mobile Objective Diagnostics of Macular Degeneration using Dark-Adapted Visual Evoked Potentials

**DOI:** 10.1101/2025.10.30.25339158

**Authors:** S. Mohammad Ali Banijamali, Craig Versek, Kameran Lashkari, Peter Bex, Srinivas Sridhar

## Abstract

**Purpose:** Delayed Dark-Adapted vision Recovery (DAR) is a known biomarker for Age-related Macular Degeneration (AMD); however, its measurement is often cumbersome for both patients and examiners. In this study, we developed NeuroVEP, a portable, wireless, and user-friendly system designed to objectively assess Dark-Adapted Visual Evoked Potentials (DAVEP).

**Methods:** NeuroVEP consists of a headset with a smartphone that delivers controlled photo-bleach and monocular pattern reversal stimuli while utilizing custom electroencephalography (EEG) electrodes and electronics to measure DAVEP. The system allows for separate analysis of the near peripheral and macular visual field of each eye, completing the test in a comfortable, single-session format (<25 minutes) without requiring subjective patient feedback.

The NeuroVEP test protocol included: (i) Mesopic luminance pattern reversal VEP for macular and peripheral regions (5 mins), (ii) Full-field photopic pattern reversal VEP (2.5 mins), (iii) Scotopic luminance DAVEP recovery post photo-bleach (up to 15 mins), measured simultaneously from both eyes.

The data were analyzed for 66 participants, divided into four cohorts: (A) Age-matched healthy controls with no ophthalmic pathologies (n=10), (B) Early-stage AMD (AREDS1) (n=19), (C) Intermediate-stage AMD (AREDS3) (n=18), (D) Advanced-stage AMD (AREDS4/5) (n=19).

Advanced signal processing and machine learning methodologies were applied to filter and process the VEP responses from the DAR segment of the experiment. 13 discriminating features were extracted from the processed signals and classified for each participant using a Bayesian statistical framework and Gaussian Mixture Model (GMM).

**Results:** The algorithm demonstrated: 86% accuracy in early-stage AMD detection (Healthy vs. Early AMD) (Sensitivity: 97%, Specificity: 65%, AUC-ROC: 0.81 and AUC-PR: 0.92) and 93% accuracy in overall AMD detection (Healthy vs. All AMD stages) (Sensitivity: 98%, Specificity: 65%, AUC-ROC: 0.82 and AUC-PR: 0.97).

**Conclusions:** We successfully developed a portable, objective user-friendly VEP system and an advanced Bayesian-GMM statistical analysis framework capable of identifying DAR deficits in AMD patients. This novel technology shows high potential for early AMD detection and could serve as a non-invasive, objective diagnostic tool for AMD screening in clinical and remote settings.

## INTRODUCTION

Age-related macular degeneration (AMD) leads to progressive central vision loss and is the leading cause of irreversible blindness in developed countries. [1] AMD impacts approximately 20 million people in the U.S. and 196 million globally. As a leading cause of severe vision impairment in older adults, its prevalence is projected to rise to 288 million worldwide by 2040. [2] According to the Age-Related Eye Disease Study (AREDS) [3], AMD progression is classified into three stages: Early, Intermediate, and Advanced.

AMD primarily affects the central retinal area responsible for high-resolution vision (the fovea and surrounding macula), while peripheral retina remains relatively intact. The loss of central vision significantly impacts quality of life and daily activities, including reading [4], face recognition [5], mobility [6], watching television [7]. Additionally, AMD has been associated with higher rates of depression [8] and an increased risk of mortality [9].

We have developed a portable, wireless system called NeuroVEP (illustrated in **Figure 1**) to objectively measure Dark-Adapted Vision Recovery (DAR) using transient Visual Evoked Potentials (VEPs) in response to pattern reversal stimuli (lasting 600-700 ms). We refer to this method as Dark-Adapted VEP (DAVEP).

**Figure 1.**
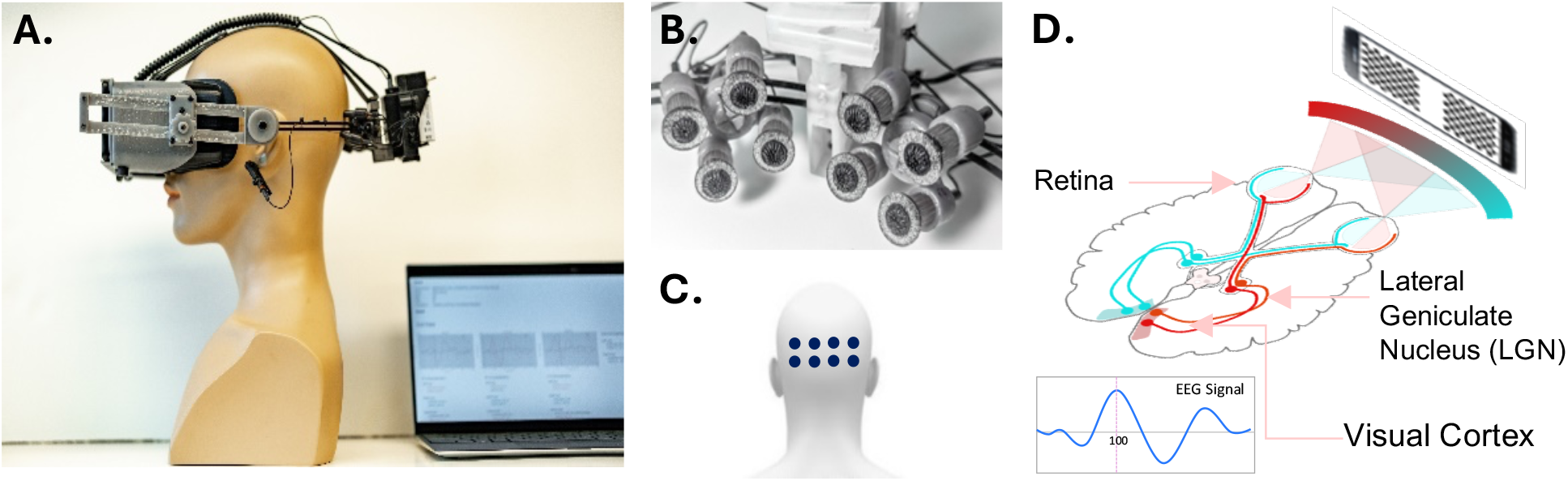
Portable wireless NeuroVEP system. A. NeuroVEP device prototype integrating NeuroVEP sensor with visual stimulus headset, B. Closeup of NeuroVEP EFEG sensor arrays, C. Location of electrodes on the scalp over the visual cortex, D. Dichoptic stimulus and typical neuroelectric response, Visual pathway. *Image in part D: Derived from (commons*.*wikimedia*.*org/wiki/File:Human_visual_pathway*.*svg)*

In this study, we assess the recovery of DAVEP responses following a photo-bleach exposure (320 cd/m^2^ for 60 seconds) over a 15-minute period using a constant scotopic luminance stimulus (5×10^−4^ cd/m^2^). The experiment was conducted on normally sighted control participants and patients across three stages of AMD (early, intermediate, and advanced).

To enhance diagnostic precision, we designed stimuli to test both eyes monocularly, targeting two distinct visual field sub-regions: the macular region (2.5–8.8° eccentricity) and the peripheral region (11.6–26.9° eccentricity), independently within the same session.

For classification, we leveraged Scientific Python, Scikit-Learn, and Statsmodels in Python to develop an automated signal processing framework and a probabilistic statistical model based on Bayesian statistics to distinguish between different stages of AMD. [10-12] The model demonstrated promising accuracy, particularly in detecting early-stage AMD.

## MATERIALS AND METHODS

Our AMD testing paradigm makes use of the NeuroVEP system, which combines scalp neuroelectric potential sensors with a smartphone in a portable wireless headset (Figure 1). In general, the system records VEPs in response to dichoptic stimuli presented on the smartphone OLED screen, which displays persistent images updated at a maximum rate of 60 frames per second. [13, 14] We used an updated prototype similar to the one that was developed in our predecessor studies [14-16]; the modifications include: a newer model smartphone (Samsung Galaxy S21 Ultra); a more ergonomic fit, with decreased weight, and better ventilation for the headset; the incorporation of a commercial eye-tracking device (Pupil Labs VR/AR add-on [12]); and a new generation of our custom hydrogel encapsulated electrodes. (see patent US 11,701,046 BS [13], example 5 for disclosure of a previous generation electrode technology. The new generation of electrodes have a similar formulation, but do not contain magnesium or lithium salts, rather potassium chloride at double physiological saline concentration is the only ionic component used. Further design details are disclosed in the section Neuro-electric Sensing below.)

### Neuro-electric Sensing

The NeuroVEP system (**Figure 1 A. – C.**) has 8 independent EEG channels, which are grouped into two positionable sensor arrays, each comprised of 4 small-diameter (10 mm) electrodes arranged in a grid with a small spacing of 2 cm (considered to be “ultra” or “very” high-density). [17] The in-house fabricated proprietary electrodes consist of a silver base functionalized with a nanoscale silver chloride coating, which is encapsulated with an environmentally stable ion conducting hydrogel that is soft yet mechanically robust. A flexible 3D-printed insulating jacket surrounds the electrode body exposing just the tip of scalp-contacting hydrogel, rimmed by a sponge-like annulus and exterior cup-shaped lip; the hydrogel tip is molded into a brush-like texture which, along with the sponge and cup, helps to hold a globule of low viscosity saline electrolyte and prevent it from leaking laterally.

The arrays slide along a track band and are coupled with a ball-socket swivel joint so that the electrode tips can rest stably against variable scalp curvatures. Tensioning racks on either side of the headset allows the contact pressure of the arrays to be precisely adjusted. The track band is attached to the headset tensioning racks with a lockable 12.5° per step incremental angle hinge. In this study, the two physical arrays have been arranged into three neighboring groups of 4 electrodes with equal spacing, referred to as LA (left array), CA (center array), and RA (right array), where CA shares the rightmost two channels of LA and the leftmost two of RA. By applying an angle of 25° (relative to nasion-inion plane) to the hinge of the track band, we target the three contiguous standard 10–20 System scalp locations O1 (LA), Oz (CA), O2 (RA).

Each of the electrodes in the array connects to its own amplifier channel, which amplifies the potential difference between it and a separate reference electrode, which is clipped to the left ear lobe of the participant. The average potential of a symmetric array (with respect to the remote reference) *V*_*avg*_ is analogous to a single EEG channel at the center of the array, albeit with lower noise. Uniquely to our system, we derive differential signals for each array called Electric-Field Encephalography (EFEG) channels. In this montage, local electric field components tangential to the scalp, *E*_*x*_ (right lateral) and *E*_*y*_ (anterior), can be estimated via *local gradients* about this central potential – *the polarity of which is independent of the remote reference location*. Using the assumptions of approximate linear variation over the small length scales of the array and the fundamental relation between the electric field and the gradient of the potential **E** = −∇*V*, we suppose that the measured potentials approximately follow a two dimensional linear form ⍰ *V*_*i*_ ≅ *– E*_*x*_*x*_*i*_ *– E*_*y*_*y*_*i*_ *+ V*_*avg*_, where index i runs over the four array electrodes, *V*_*i*_ stands for the ith sensor’s potential, and (*x*_*i*_,*y*_*i*_) is its spatial coordinates; hence, the determination of the electric field components tangential to the scalp can be treated as the slope parameters in a plane fitting problem (with zero intercept after subtracting *V*_*avg*_) for which we use the ordinary least squares solution. Some of the interesting properties of the EFEG montage have been evaluated in our previous publications [18, 19].

The recordings for each EEG channel are sampled at a rate of 1000 samples per second with a resolution of 24 bits and a gain factor of 24, and the data is saved, unfiltered, in a binary file format. Before subsequent data processing, each channel is prefiltered using a Stationary Wavelet Transform (SWT) baseline removal [20] with an effective cutoff frequency of 0.5 Hz; we find that this process preserves the time domain characteristics of VEP responses while greatly reducing small motion artifacts and electrode polarization artifacts that might otherwise create longer duration transient distortions (impulse/step response of filter) using standard digital filtering techniques. Any additional filtering steps will be described in the “Results” sections below.

### Paradigm Schedule Overview

During the same testing session, participants’ eyes are tested monocularly through a sequence of high contrast dartboard pattern-reversal visual stimuli:

1. **Mesopic-Level Pattern Reversal VEP (“mesVEP”):** A 5-minute test using a 0.2 cd/m^2^ green light stimulus, targeting independent macular and peripheral visual field regions (detailed in the “Visual Stimuli” section, Figure 3).
2. Photopic-Level Pattern Reversal VEP (“photVEP”): A 2.5-minute test using a 16 cd/m^2^ white light stimulus, covering a full-field region (2.5–26.9° eccentricity).
3. **Photobleaching Phase:** – A 60-second exposure to a 320 cd/m^2^ full-intensity white screen, covering up to 45° eccentricity.
4. **Scotopic-Level Dark Adapted VEP (“DAVEP”):** – A 15-minute test using a 5×10^−4^ cd/m^2^ green light stimulus to measure dark-adapted vision recovery, utilizing the same macular and peripheral regions as in the “mesVEP” test (Figure 3).

See **Figure 2** for an overview of the testing sequence.

**Figure 2.**
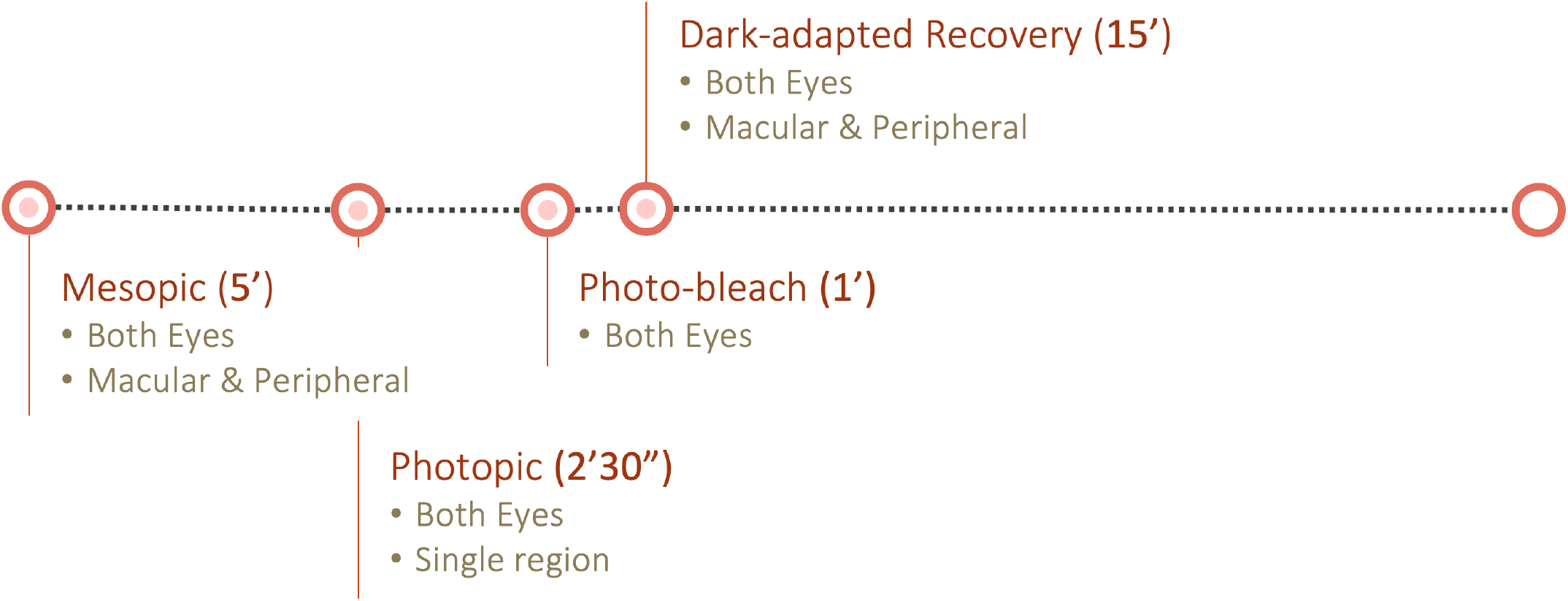
Paradigm Overview.

**Figure 3.**
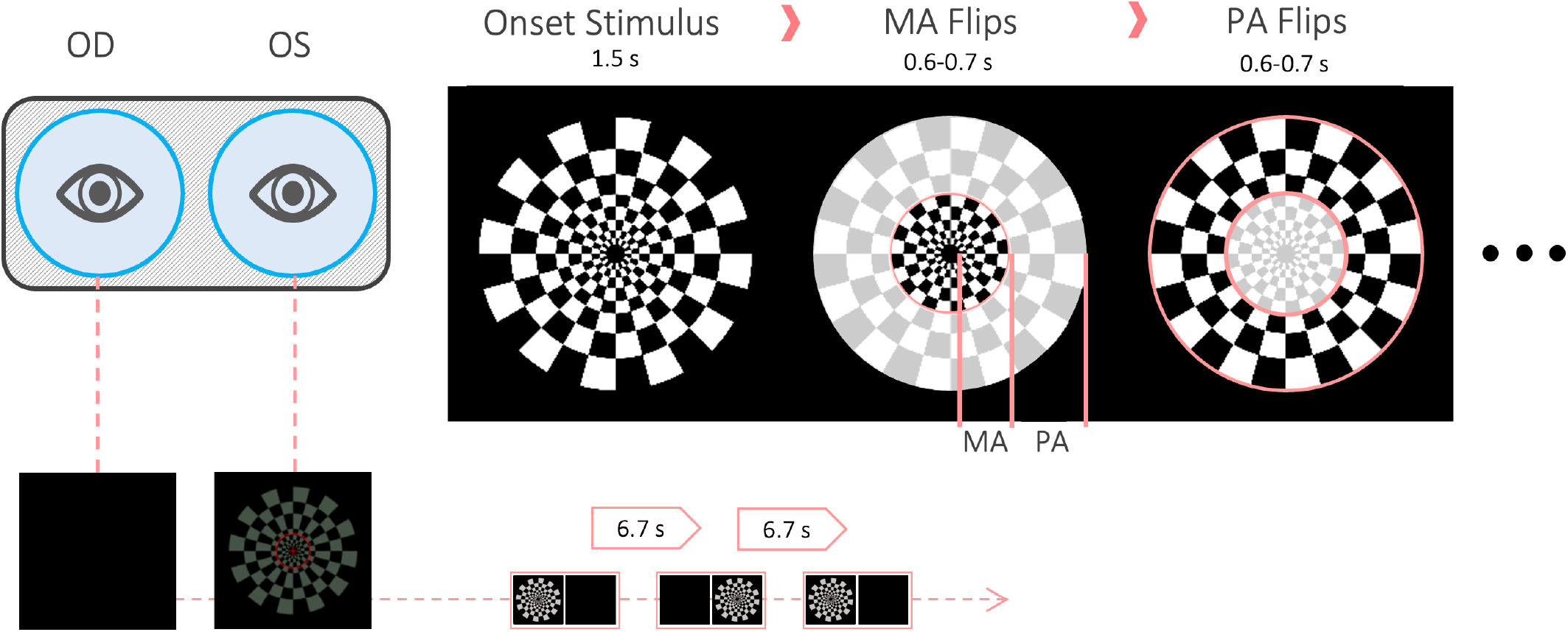
The test stimuli for the DAVEP paradigm consist of a log polar dartboard pattern which is divided into the “Macular Annulus” (MA, 2.5–8.8° ecc.) and “Peripheral Annulus” (PA, 11.6– 26.9° ecc.) visual field regions whose check pattern reverses independently. Stimuli are presented monocularly (to each eye with the opposite eye in complete darkness): after an unmeasured “onset” period of 1.5s, the pattern reverses 4 times for each visual field region with a step of 0.6–0.7 s (randomized); the sequence lasts about 6.7 s before switching to the other eye. See the text for an exact description of the sequence.

We did not use the Mesopic and Photopic tests for participant classification. Instead, these tests were included to identify individuals with severe visual field dysfunctions that would render them ineligible for the study. However, no participants with such disabilities were observed in our study.

### Visual Stimuli

A dartboard-patterned polar checkerboard reversal was used to evaluate the eye-brain system’s response to mesopic, photopic, and scotopic luminance stimuli, following the schedule outlined in the previous section.

The stimuli were displayed on the OLED screen of a Samsung Galaxy S21 Ultra smartphone, integrated into a custom VR head-mount with optics similar to those of the Google VR Daydream View (first version, 2016). Since OLED displays emit no light from black pixels, the contrast was practically infinite and beyond participants’ perceptual range, provided that reflected light was minimal. To ensure this, all experiments were conducted in a windowless room with the lights turned off.

The stimuli patterns were generated as high-resolution pixel maps (2048x2048) using custom Python programs, then were loaded into a custom app using the Google Cardboard VR Android SDK utilizing OpenGLES2 texture rendering. The dartboard pattern (Figure 3) was divided into two distinct regions for the “mesVEP” and “DAVEP” tests:

- Macular Annulus (MA): Covers most of the perifoveal macula (2.5–8.8° eccentricity), excluding the fovea.
- Peripheral Annulus (PA): Covers the near peripheral region (11.6–26.9° eccentricity) of each eye.

The annular dartboard patterns were radially scaled following the cortical magnification rule, [21] where the radial length R(E) at eccentricity E (degrees of visual angle) scales by R(E)/R(0) = 0.27E + 1, with a base check size of R(0) = 1° of visual field subtended. The stimuli were either white-colored (for “photVEP”; uniform pixel values for the three RGB display channels) or solid green-colored (for “mesVEP” and “DAVEP”; single channel on RGB display at wavelength 532 ± 20 nm, closest to the rod photoreceptor sensitivity centroid of 498 nm), although they appear virtually colorless at scotopic intensity levels. The foveal region of 0–2.5° was excluded to minimize the contribution of cone photoreceptors which have the highest density there.

Unlike in our predecessor study [14], in which the MA region spanned 2.6-16°, we did not extend it far beyond the physiologically defined macula at 6°; this limiting is intended to improve sensitivity to earlier stages of AMD, but it is also expected to change the quality of the macular response differentiating it further from the peripheral and perhaps lowering the SNR due to the lower amount of targeted rod photoreceptor cells (rod density increases from zero just outside the fovea to a peak at 18° eccentricity, but falls off more slowly in the periphery [22]).

A neutral density filter of 10 stops (log base 2 units) was inserted immediately after the completion of the photo-bleach to achieve scotopic luminance of 5×10^−4^ cd/m^2^ which was not possible using only the native display range; this stimulus level was chosen in the upper scotopic regime to limit the expected response amplitude recovery times for normal participants below the 15 min DAVEP test duration. A mid-mesopic intensity red fixation cross, spanning the foveal region, and a thin ring at 10° eccentricity were overlaid on the stimuli to help participants, especially those with central vision loss, maintain fixation at the center.

Both the “mesVEP” and “DAVEP” tests alternate regional pattern reversals according to two fixed sequences: a) S, M, P, M, P, P, M, P, M; b) S, P,M,P,M,M,P,M,P, where S is the starting image (1.5 s onset, not measured), and M and P are macular and peripheral region reversals, respectively (at an interval of 0.6 s plus a random jitter from 0 to 0.1 s). The stimuli are presented monocularly, such that each sequence is cycled between the left L, then the right R eye – lasting ∼6.2 s in each eye, where the opposite eye is in complete darkness – the period to cycle through all four sequences L.a, R.a, L.b, R.b lasts for an average duration of 27 s. The sets of stimuli are broken into 2.5-minute phases which include 5 repetitions of the full cycle, thus averaging 135 s; the remaining ∼15 s of each phase is announced (with a synthetic voice, text-to-speech) as a short break period in which participants are permitted to shift positions and blink as desired for comfort. In the DAVEP test the remaining time is announced every 5 minutes (2 phases).

### Participant Pool

All studies were approved by the Northeastern University Institutional Review Board (IRB) (protocol #17-09-01) and conducted in accordance with the Declaration of Helsinki.

Study participants with AMD and control participants were referred by their vitreoretinal specialist at Advanced Eye Centers (Dartmouth, MA). Participants underwent dilated fundus exam and optical coherence tomography (OCT) imaging and were classified according to the AREDS study [3]. Also, some controls were referred by other ophthalmologists, recruited through personal contacts, or enrolled via the PsyLink course credit granting experiment participation system at Northeastern University.

Participants were required to be at least 18 years old and provided informed consent after a detailed explanation of their role in the study. Gender was not a selection criterion. Control participants were age-matched to ensure a more precise comparison with other participant groups. The total number of participants tested was 98 distributed as follows: Controls 26, Early (AREDS 1) 24, Intermediate (AREDS 3) 24, Advanced (AREDS 4/5) 24. Data from 32 participants was excluded for the following reasons:

- Corrupt and un-analyzable test data files: This affected 20 cases across four categories.
- High noise or elevated alpha wave levels, resulting in a very poor signal-to-noise ratio (SNR) in VEP responses, EFEG responses, or all responses. This issue was observed in 12 control participants.

The poor SNR in excluded control participants may be attributed to:

- Limited stimulus sensitivity for older participants with impaired vision.
- Potential confounders, such as presbyopia.
- Participant-related factors, including lack of focus, attention, drowsiness, eye closures, or excessive movement during the test.

The characteristics of the participant pool of 66 included in the final data analysis are described in **Table 1**.

**Table 1.**
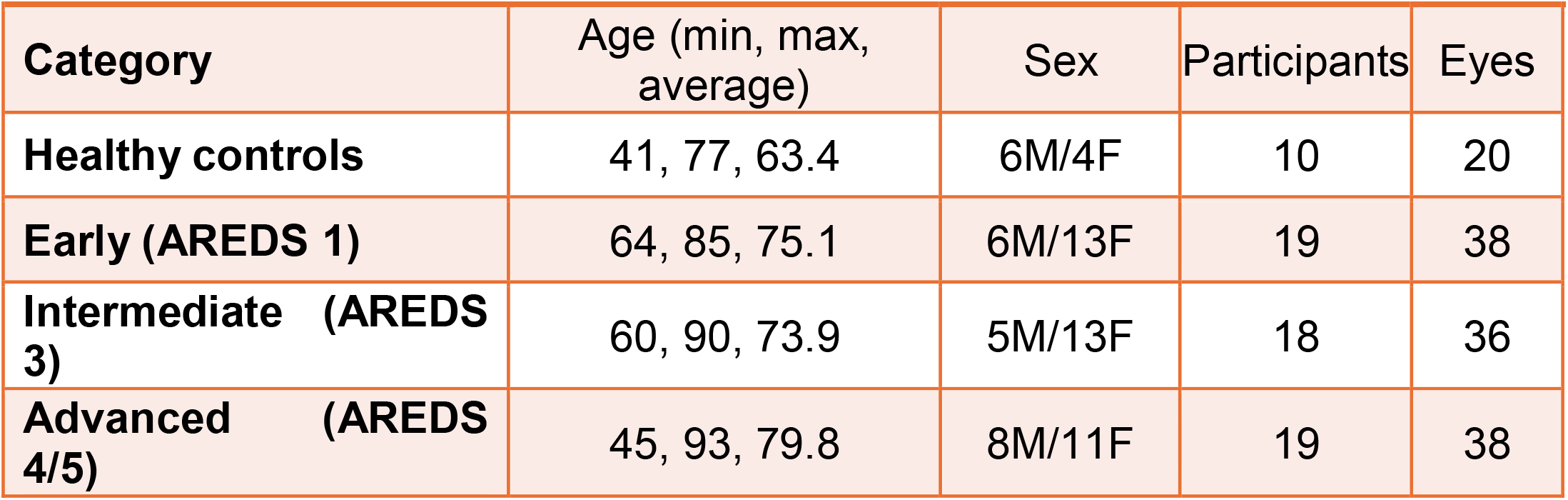
Participant pool characteristics included in data analysis.

| Category | Age (min, max, average) | Sex | Participants | Eyes |
| --- | --- | --- | --- | --- |
| Healthy controls | 41, 77, 63.4 | 6M/4F | 10 | 20 |
| Early (AREDS 1) | 64, 85, 75.1 | 6M/13F | 19 | 38 |
| Intermediate (AREDS 3) | 60, 90, 73.9 | 5M/13F | 18 | 36 |
| Advanced (AREDS 4/5) | 45, 93, 79.8 | 8M/11F | 19 | 38 |

After the tests, all participants were required to evaluate their experience with the test and the headset’s hardware. The questionnaire results are summarized in **Table 2** based on percentage of responses in each category.

**Table 2.** Results of usability of the device questionnaire.

| Questions |  | Percentage |
| --- | --- | --- |
| Please rate your level of comfort with the headset hardware. | Very Comfortable | 13 |
|  | Comfortable | 62 |
|  | Uncomfortable | 25 |
|  | Very Uncomfortable | 0 |
| Did you experience any eye strain with the device? |  | 3 (Yes), 97 (No) |
| Are you comfortable with the length of the experiment? |  | 95 (Yes), 5 (No) |
| Would you recommend this test to someone else? |  | 99 (Yes), 1 (No) |

### Clinical tests

Visual acuity was assessed using the ETDRS (Early Treatment Diabetic Retinopathy Study) chart under both high and low light conditions, a methodology adapted from the Comparison of Age-Related Macular Degeneration Treatments Trials (CATT) protocols [23, 24]. OCT measurements were performed using the Spectralis OCT/SLO (Heidelberg Engineering, Germany).

### Test Administration

Most clinical participants were unfamiliar with EEG/VEP but were instructed to minimize body movements, eye closures, and talking during the recording. However, they were explicitly allowed to blink freely and adjust their posture during ∼15-second break periods for comfort. Participants were required to wear N95 masks throughout the test, and the device was thoroughly disinfected with alcohol after each test.

No pupil-dilating drugs were administered, and no artificial pupil restrictions were used in any of the reported tests. We had participants remove any eyeglasses before placing the device over their heads. After securing the head-mounted display and neuroelectric sensors, the experimenters applied a skin-safe saline electrolyte with gentle swabbing around the electrode/scalp contact area, while monitoring EEG channel impedances to ensure they remained below 100 kΩ—a threshold previously shown to be sufficient for high-quality measurements using the NeuroVEP system. [19] No abrasive preparation compounds, conductive pastes, or gels were applied to the scalp. Additionally, any bulky jewelry or headwear that could interfere with the electrodes or mounting hardware was removed before the experiment.

The tests caused minimal discomfort but required participants to maintain alertness for extended periods. To help manage fatigue, the tests were broken into shorter 2.5-minutes segments with brief pauses (15–30 seconds), during which automated voice instructions and remaining time announcements were provided.

Due to the repetitive nature of the visual stimuli and low light conditions during the DAVEP test, there was a concern about boredom and microsleeps. To mitigate this, participants were encouraged to listen to their favorite music or a podcast throughout the testing session.

### Fixation Compliance

Adequate central fixation on the stimuli by participants is a concern for restricted eccentricity pattern VEP paradigms such as DAVEP, especially for highly impaired AMD patients with central vision loss: as described in the section “Visual Stimuli”, we provide an extended fixation target at 10° eccentricity to increase visibility for central vision impaired participants; additionally, response metrics are based on averages of many trials, so the effects of temporary, infrequent loss of fixation are mitigated. We used *in situ* eye-camera monitoring provided by the Pupil Labs VR/AR addon, which allowed the experimenter to verbally correct obvious issues such as excessive eye-closure or skewed fixation; however, we did not record eye-tracking information since the data stream proved to be too unreliable with frequent detection dropouts.

### DAVEP Processing Pipeline

#### I Signal Processing, Data Cleaning, and Filtering

The NeuroVEP system ensures precise synchronization of video frame events by using a phototransistor detector to monitor a hidden screen patch dedicated to transmitting this signal. These stimulus event markers are then utilized in data processing to accurately align and average neuroelectric responses from the exact onset of each stimulus presentation.

Epochs are defined from the stimulus onset up to 600 ms, ensuring the response has decayed below the noise floor. Data is grouped separately by left (L) or right (R) eye and macular (M) or peripheral (P) regions (S stimulus onsets are not processed).

To enhance data quality, an unsupervised machine learning outlier rejection algorithm (Scikit-Learn’s Isolation Forest model) is applied to variances of trial signals with and without 1 Hz cutoff Bessel IIR high-pass filtering. This method identifies and excludes trials with outlier variances, typically caused by movement artifacts. Only a small percentage of trials (up to 10%) are removed, preserving the integrity of the windowed average.

#### II Spatiotemporal Data Preparation

Figure 4. shows the signal processing and data classification pipeline. To analyze the DAR data, we divided the recovery period into two segments: the first and the second half of the test period. We then averaged all trials within each half, resulting in two individual signals representing the first and second segments of the recovery. Averaging the trials in these two segments helps to eliminate random noise and alpha waves. We applied this procedure to the following signals:

1. EEG, the average of all EEG potentials from the 8 channels,
2. E_y_, the E_y_ component of EFEG,
3. E_x_(R-L), the difference between the E_x_ component of EFEG in the right and left hemispheres.

**Figure 4.**
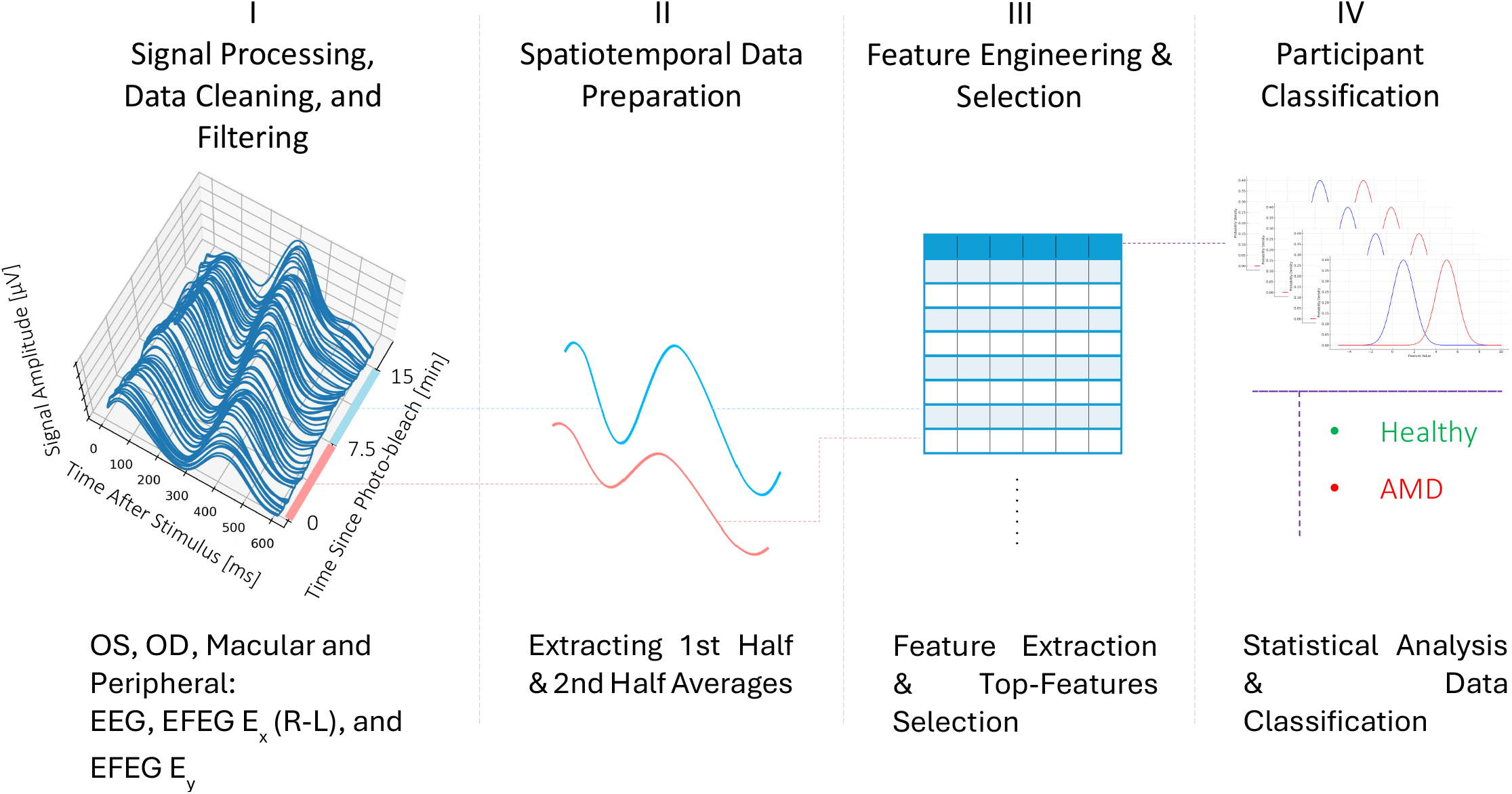
Signal processing and classification pipeline for the DAR portion of the experiment.

These signals were collected from both the macular and peripheral regions of the visual field. Together, a total of 12 distinct signals were analyzed per eye for each participant.

By overlaying these signals for each participant group (healthy (n=10, 20 eyes), early AMD (n= 19), intermediate AMD (n=18), and advanced AMD (n=19)), we can visually discern differences between the groups. **Figure 5** highlights the differences in signal responses between a healthy participant and a participant with advanced AMD. In each plot, the first-half DAR average responses are overlaid with the second-half average responses to visualize progression over time. The recovery dynamics are particularly evident in the healthy participant, where a clear distinction between the two halves indicates a strong adaptive response compared to the advanced AMD case. To quantify the differences, we extracted a set of features from these signals and examined the variations in the distributions of values for each parameter across the participant categories.

**Figure 5.**
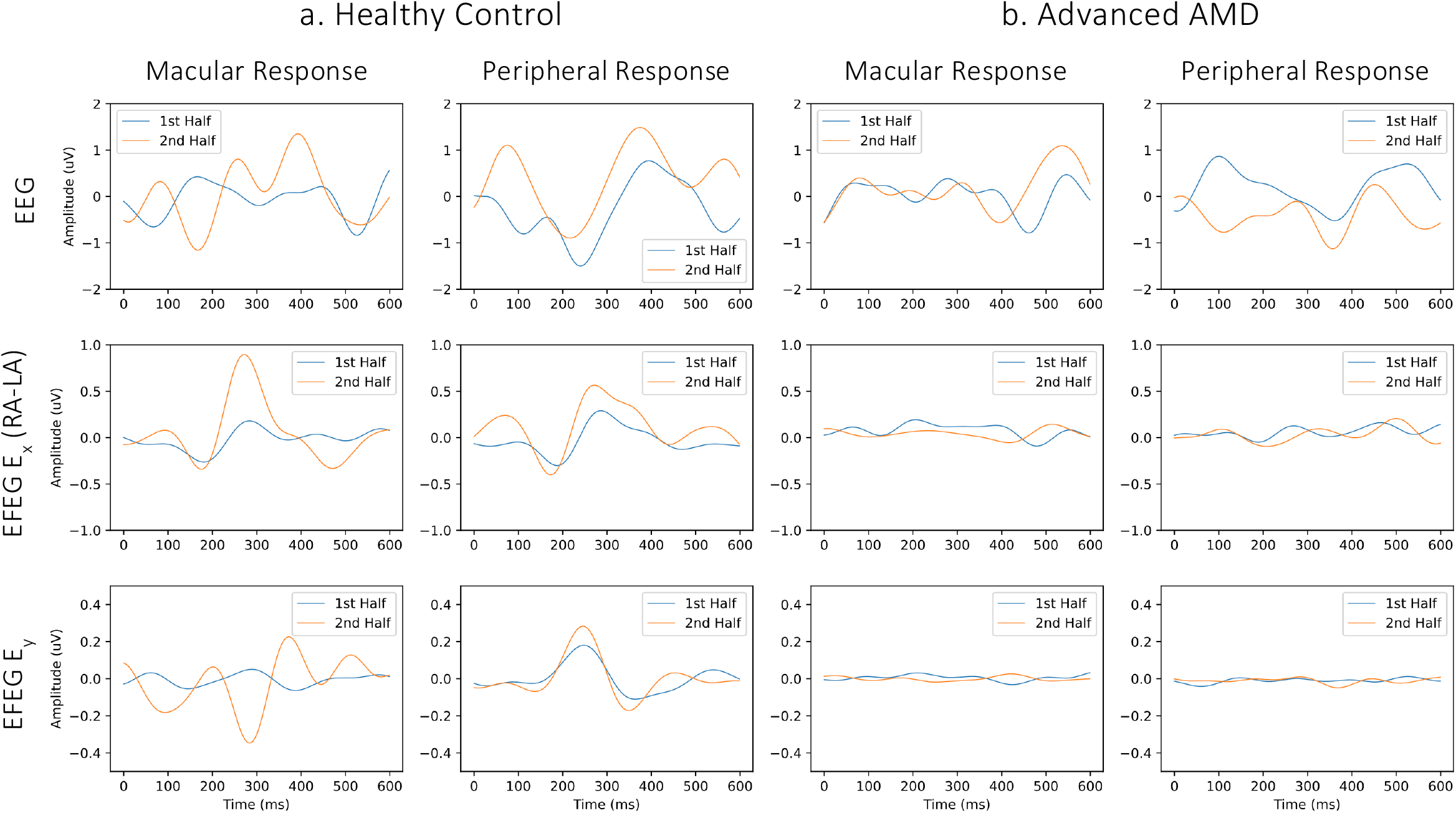
EEG, EFEG: E_x_ (Right Array – Left Array) and E_y_macular and peripheral responses for a. healthy subject and b. advanced AMD subject. Each plot shows average response of first vs. second half of the DAR portion of the test.

#### III Feature Engineering and Selection

Our primary focus in this study was on the early detection of AMD, so we paid particular attention to the differences between the healthy participants and the early AMD group. We used the two-sample Kolmogorov-Smirnov test to compare the distributions and identify features that effectively distinguish between these two categories. The null hypothesis of the test is that the two distributions are identical, F(x)=G(x) for all x; the alternative is that they are not identical. Based on the p-values from the test, we selected the top-performing features, identifying 13 features with **p-values less than 0.003** from macular and peripheral responses that can reliably differentiate between healthy and early AMD participant groups. The features are presented in **Table 3**, and their boxplot for different stages of AMD is shown in **Figure 6**.

**Table 3.** List of top performing features and their definition

|  |
| --- |
| m_ey_c_h2_signal std<br>Macular – Ey (Center Array) – 2<sup>nd</sup> Half – Standard deviation of signal region of the response |
| p_ey_c_h1_signal std<br>Peripheral – Ey (Center Array) – 1<sup>st</sup> Half – Standard deviation of signal region of the response |
| p_ey_c_h1_absolute_maximum<br>Peripheral – Ey (Center Array) – 1<sup>st</sup> Half – Absolute maximum of the response |
| p_ey_c_h2_signal std<br>Peripheral – Ey (Center Array) – 2<sup>nd</sup> Half – Standard deviation of signal region of the response |
| p_ey_c_h2_absolute_maximum<br>Peripheral – Ey (Center Array) – 2<sup>nd</sup> Half – Absolute maximum of the response |
| m_ex_ra_la_h1_snr<br>Macular – Ex (Right Array – Left Array) – 1<sup>st</sup> Half – SNR |
| m_ex_ra_la_h2_signal std<br>Macular – Ex (Right Array – Left Array) – 2<sup>nd</sup> Half – Standard deviation of signal region of the response |
| m_ex_ra_la_h2_Lempel_ziv_complexity_bins_100<br>Macular – Ex (Right Array – Left Array) – 2<sup>nd</sup> Half – Lempel-Ziv complexity, bins=100 |
| m_ex_ra_la_h2_Lempel_ziv_complexity_bins_10<br>Macular – Ex (Right Array – Left Array) – 2<sup>nd</sup> Half – Lempel-Ziv complexity, bins=10 |
| p_ex_ra_la_h1_Lempel_ziv_complexity_bins_100<br>Peripheral – Ex (Right Array – Left Array) – 1<sup>st</sup> Half – Lempel-Ziv complexity, bins=100 |
| p_ex_ra_la_h1_Lempel_ziv_complexity_bins_10<br>Peripheral – Ex (Right Array – Left Array) – 1<sup>st</sup> Half – Lempel-Ziv complexity, bins=10 |
| p_ex_ra_la_h1_binned_entropy_max_bins_10<br>Peripheral – Ex (Right Array – Left Array) – 1<sup>st</sup> Half – Binned entropy, max bins=10 |
| P_dar_h2_signal std<br>Peripheral – DAR– 2<sup>nd</sup> Half – Standard deviation of signal region of the response |

**Figure 6.**
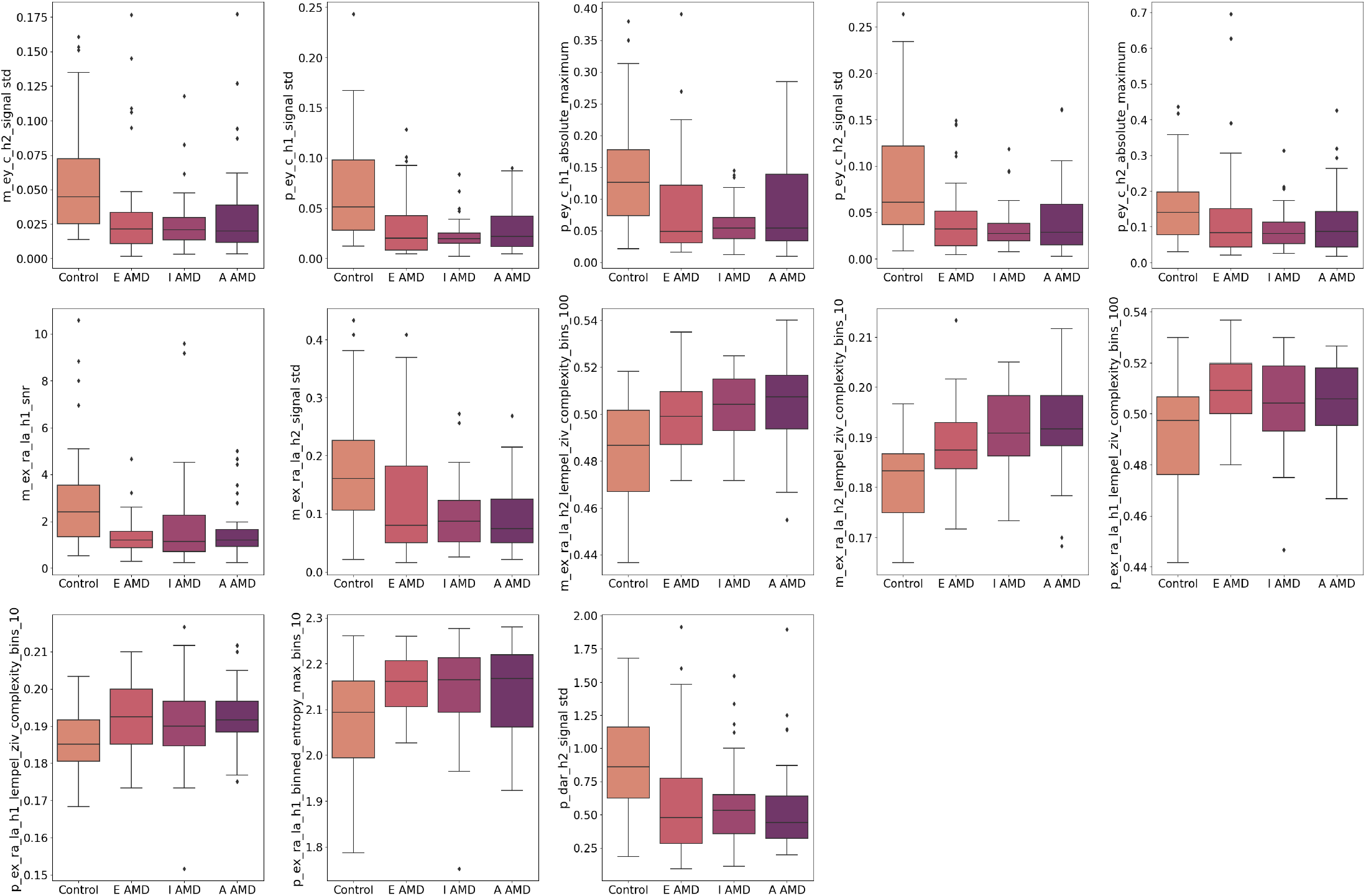
Boxplots showing the distribution of top performing features for healthy controls vs. 3 stages of AMD.

#### IV Participant Classification

This data could be used for training a machine learning algorithm to classify participants. However, due to insufficient data for training and validating a machine learning classification algorithm, we opted to develop a statistical framework for participant classification. This framework addresses a binary classification problem, where participants are categorized as either healthy or AMD. We used the features from the healthy group of participants for the healthy category and the feature set from the early AMD category for the AMD group. To evaluate the early detection capability of our method, we made the classification task more challenging by setting the criteria for classification as an AMD participant based on the features of the early AMD group. To determine the probability of a participant belonging to the AMD group, given the two distributions, we utilized Bayes’ Theorem as follows:

For each single feature (component), we have probability density functions (PDFs) *f*_*H*_(*x*) and *f*_*A*_ (*x*) for healthy control participants *p*(*H*) and early AMD *p*(*A*), respectively. Assuming prior probabilities *p*(*H*) and *p*(*A*), the posterior probability of observing a data point *x* belonging to class *A* is calculated using Bayes’ Theorem:

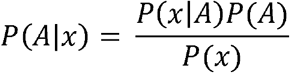

The term *p(x)* is the total probability of observing x, computed using the law of total probability:

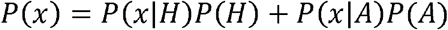

Here, *p*(*H*) and *p*(*A*) are the initial probabilities of the classes before observing any data, representing our prior belief about the likelihood of each class based on prior knowledge or assumptions. We assume these probabilities to be 0.5 each.

This calculation is for a single feature. However as discussed above, we have identified 13 top-performing features that can distinguish between healthy participants and the Early AMD class. When multiple distributions exist for each class, it suggests that each class comprises several subgroups, each with its own distribution. This scenario can be addressed using a mixture model, such as a Gaussian Mixture Model (GMM), where each class is represented as a combination of multiple distributions.

In the context of Bayesian classification, we still use Bayes’ Theorem, but the likelihoods *p*(*x*|*A*) or *p*(*x*|*H*)for the Healthy or Early AMD class are now weighted sums of the likelihoods of the individual distributions (components) constituting each class.

Based on this approach, again we have two classes: healthy (H) and early AMD (A). Each class is represented by *k* = 13 components, where each component corresponds to a distribution for a feature *θ*_A/H,i_ with *i* = 1, …, *k*. Each component is assigned a weight. Since all features were selected based on a similar criterion (with p-value < 0.003) and the p-values were very close, we assumed equal weights for all components. The weights are defined such that:

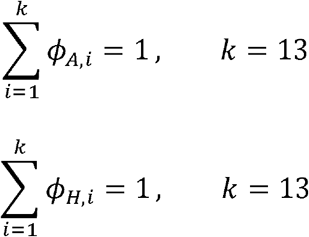

Next, the likelihood *p(x*|*A)*, is calculated as the weighted sum of the likelihoods of all its components:

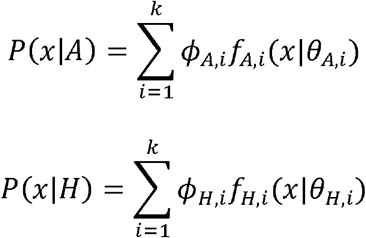

Where 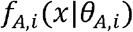 is the PDF of the i-th component of class A (AMD)).

Finally, given the prior probabilities *p*(*A*) and *p*(*H*) (both assumed to be 0.5), the posterior probability based on Bayes’ theorem will be:

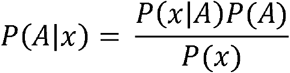

Where *p(x)* is the total probability of observing x and is calculated by:

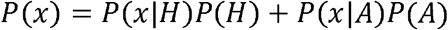

Using this method, we calculated the probability of each eye/ participant having AMD. We then converted these probabilities into classes by defining probabilities below 0.5 as healthy (class 0) and those above 0.5 as having AMD (class 1). Subsequently, we evaluated the classification task by calculating sensitivity, specificity, and accuracy metrics (see Results).

All analyses were conducted using the Python programming language, utilizing scientific libraries such as SciPy, Scikit-learn, and Statsmodels. [10, 12, 25]

## RESULTS

### Classification Results

We conducted the classification task twice: first, to distinguish all AMD stages from the control group, and second, to assess the model’s early detection capability by focusing specifically on early-stage AMD. For each case, we calculated accuracy, sensitivity, and specificity. Additionally, given the class imbalance, particularly in the control vs. all AMD stages scenario, we also calculated the Area Under the Receiver Operating Characteristic Curve (AUC-ROC) and Area Under the Precision-Recall Curve (AUC-PR) to provide a more comprehensive evaluation of the model’s performance. The results are summarized in **Table 4**.

**Table 4.** Performance metrics for two classification scenarios.

|  | Accuracy | Sensitivity | Specificity | AUC<br>ROC | AUC<br>PR |
| --- | --- | --- | --- | --- | --- |
| All Stages of<br>AMD | 0.93 | 0.98 | 0.65 | 0.82 | 0.97 |
| Early-Stage AMD | 0.86 | 0.97 | 0.65 | 0.81 | 0.92 |

### Effect Size and Statistical Power Analysis

Using the calculated classification probabilities, we performed statistical power and effect size analysis for two classification scenarios: control vs. all stages of AMD and control vs. early-stage AMD.

First, we determined the effect size using Cohen’s d, which resulted in 2.05 and 1.70, respectively. To account for the small sample size, we applied Hedges’ g correction, yielding effect sizes of 2.04 and 1.68 for the two classification problems.

Next, using the Hedges’ g effect sizes, we calculated the statistical power based on a two-sample t-test with a significance level of α = 0.05, obtaining values of 1.00 for control vs. all AMD and 0.99 for control vs. early AMD, indicating strong statistical significance. (**Table 5**)

**Table 5.** Power analysis and statistical significance.

| **Table 5.** Power analysis and statistical significance. |  |  |  |  |
| --- | --- | --- | --- | --- |
|  | **Cohen's d**<br>**Effect Size** | **Hedges' g**<br>**Effect Size** | **Significance ($\alpha$)** | **Statistical**<br>**Power** |
| **All Stages of AMD** | 2.05 | 2.04 | 0.05 | 1 |
| **Early-Stage AMD** | 1.70 | 1.68 | 0.05 | 0.99 |

## DISCUSSION

### Dark-adapted visual evoked potentials (DAVEP) using NeuroVEP can objectively detect the rod-mediated deficits characteristic of age-related macular degeneration (AMD)

After photobleach, a robust positive-going scotopic VEP peak at latencies from 200–300 ms in healthy eyes emerged over the 15 min recovery time as judged by comparing first and second halves of dark adaptation recovery trials, while AMD eyes exhibited comparatively reduced amplitude and delayed recovery [14]. This aligns with prior psychophysical research showing prolonged rod-mediated dark adaptation recovery as an early hallmark of AMD [26, 27]. By employing a smartphone-based headset (NeuroVEP) and high-density scalp electrodes, we captured scotopic VEP signals in both eyes and 2 locations macular and peripheral under 25 minutes without any patient response, thus streamlining the often-cumbersome process of dark adaptation testing. Our findings emphasize that DAVEP offers an objective biomarker. The DAVEP protocol does not rely on subjective patient responses, making it potentially advantageous for large-scale screenings or for individuals who struggle with conventional psychophysical methods.

Conventional psychophysical dark adaptometry is widely recognized as a sensitive means of detecting early AMD-related rod dysfunction [26, 27]. Tools like the AdaptDx [27] can distinguish early AMD from normal aging but often require prolonged testing and active patient feedback. By contrast, our VEP-based test provides a neurophysiological correlate of rod recovery without participant input. Early investigations into scotopic VEPs, such as those by Airas and Petersen, highlighted the feasibility of objectively measuring dark adaptation but were hampered by poor signal quality and artifacts [28]. We build on those foundations with advanced hardware and sophisticated signal processing and statistical analysis methods. Our results also accord with electroretinography (ERG) studies indicating diminished scotopic sensitivity in AMD, yet VEP recordings are simpler to administer and focus on cortical signals that reflect higher-level integration [14]. Compared to prior objective methods (e.g., multifocal ERG), the NeuroVEP headset is more comfortable and portable, showing promise for routine or remote testing. While some investigators have attempted VR-based dark adaptation platforms [29], our approach is distinctive in using a targeted scotopic VEP paradigm to measure cortical re-engagement of rod pathways. These observations collectively underscore that scotopic functional assessment can detect AMD at stages where visual acuity remains normal.

### Statistical analysis yielded a robust set of 13 significant features for DAVEP analysis

Traditional analysis of photopic VEP responses measuring just two parameters latency and amplitude are unsuitable for the DAR VEP signal analysis. For classification of our results, we analyzed multiple signals from different scalp locations and their combinations. Specifically, we considered the E_y_ component of EFEG at the central array location, the E_x_ component of EFEG at the right and left array locations, as well as the averaged EEG response. Incorporating various signals provided richer spatial information about cortical activation sources.

From these signals, using a Kolmogorov-Smirnov test, we extracted a robust set of 13 discriminating features with p < 0.003, either based on wave amplitude and shape (SD, absolute maximum, SNR) or wave complexity (Lempel-Ziv complexity, entropy). Additionally, temporal segmentation of the recovery process allowed us to capture the dynamic nature of dark adaptation. The key features are summarized below:

1. <u>Standard Deviation (SD) of the Signal Region:</u> Several top features measured the SD within a specific time window (175–450 ms) of the averaged VEP waveform. A higher SD often indicates a stronger or more variable cortical response. For healthy controls, a robust re-emergence of the scotopic VEP, especially in the second half of dark adaptation, yields larger fluctuations and hence increased SD. In contrast, AMD participants often display flatter waveforms or delayed peaks, resulting in a reduced SD. Notably, separate “SD” features for macular (m_ey_c_h2_signal_std) and peripheral (p_ey_c_h2_signal_std) regions highlight the fact that AMD primarily affects the macular DAVEP amplitude, although peripheral signals can also offer comparative insight.
2. <u>Absolute Maximum Amplitude:</u> Features capturing the maximum amplitude in the first or second half of the waveform (e.g., p_ey_c_h1_absolute_maximum) reflect the peak cortical response level. Healthy eyes typically show a distinct scotopic peak, whereas in AMD this peak is diminished, delayed, or absent.
3. <u>Signal-to-Noise Ratio (SNR):</u> The SNR feature (m_ex_ra_la_h1_snr) quantifies how much the DAVEP response rises above the background noise. Macular SNR in early recovery phases (first half) can be particularly telling in AMD, as these patients fail to generate a clear response shortly after bleach, in contrast to healthy controls.
4. <u>Signal Complexity (Lempel-Ziv Complexity and Binned Entropy):</u> Lempel-Ziv complexity (e.g., m_ex_ra_la_h2_Lempel_ziv_complexity_bins_100) and binned entropy (p_ex_ra_la_h1_binned_entropy_max_bins_10) capture the degree of morphological intricacy in the waveform. A healthy, well-recovered scotopic VEP often has a characteristic peak structure, whereas AMD waveforms can be irregularly flat or feature multiple sub-peaks caused by incomplete rod pathway recovery. These complexity metrics proved sensitive to such subtle shape differences, allowing the Bayesian-GMM classifier to tease apart nuanced AMD phenotypes from normal variability.
5. <u>First-Half vs. Second-Half Parameters:</u> Finally, splitting the recovery into two temporal segments was crucial. Early AMD deficits often manifest as a reduced or delayed response in the first half of testing, but as time progresses, some partial recovery may occur. Thus, incorporating features that distinguish “first half” vs. “second half” responses (e.g., p_ey_c_h1 vs. p_ey_c_h2) significantly boosted classification power.

### VEP features analyzed with a Bayesian GMM can effectively distinguish both early and advanced AMD from healthy aging eyes

We utilized Bayesian Gaussian Mixture Model (GMM) for our data analysis framework to classify eyes as AMD-affected or normal based on their DAVEP features. The decision to use a Bayesian-GMM was driven by its ability to model the underlying probabilistic distribution of our metrics while accounting for inter-participant variability and uncertainty. Essentially, the GMM treated the population of responses as a mixture of two statistical clusters, presumed to represent “normal” adaptation and “impaired” adaptation (Early AMD, or AMD at any stage).

The objective was twofold: (1) distinguish all stages of AMD from healthy controls and (2) assess the model’s ability to detect early-stage AMD. The analysis yielded two sets of performance metrics: accuracy, sensitivity, specificity, area under the ROC curve (AUC-ROC), and area under the precision–recall curve (AUC-PR). In both classification tasks, sensitivity was exceptionally high, whereas specificity was more moderate. Further interpretation was provided through effect size estimation and statistical power analysis.

In the first scenario, classifying all stages of AMD versus controls, the model achieved an accuracy of 93%, with sensitivity at 98% and specificity at 65%. These values indicate that the classifier correctly identified nearly all AMD-affected eyes (including advanced forms), but it also produced more false positives among healthy controls than would be ideal. In screening applications, however, a modest specificity can be acceptable if the disease in question has serious consequences and would require further confirmatory diagnostics to validate a positive test result. The second classification task, focused on early-stage AMD, had an accuracy of 86%, with sensitivity at 97% and specificity again at 65%. Although overall accuracy dropped slightly when targeting early disease, the high sensitivity highlights the model’s ability to detect subtle changes in VEP data that can manifest during the earliest phases of AMD.

These results were further explored through the model’s receiver operating characteristic (ROC) curves, where the area under the curve was 0.82 for all AMD stages and 0.81 for early-stage AMD. Both figures exceed 0.80, suggesting excellent discrimination between diseased and normal eyes across various decision thresholds. Notably, because the classes were imbalanced, particularly in the all-AMD dataset, the area under the precision-recall (PR) curve was also calculated. The AUC-PR values were 0.97 for all AMD and 0.92 for early AMD, highlighting the classifier’s strong ability to balance precision and recall, even with the uneven class distribution. The near-perfect AUC-PR in the all-AMD comparison underscores the model’s capacity to detect true positives reliably while minimizing false positives in a screening context where the majority of individuals might be healthy.

Beyond these core performance metrics, the study investigated the degree to which the VEP-based feature distributions differed between control and AMD groups. Effect sizes, measured by Cohen’s d, were 2.05 (all AMD) and 1.70 (early AMD), both of which are considered “very large” according to standard benchmarks. Even after applying Hedges’ g for small-sample bias correction, the effect sizes (2.04 and 1.68) remained almost unchanged. Such large effect sizes suggest that the classifier’s outputs for AMD and healthy eyes are well-separated, indicating a strong physiological distinction in VEP patterns associated with AMD pathology. In addition, power analyses based on these effect sizes showed that the study had near-100% statistical power to detect true differences, making it highly unlikely that the observed separation of AMD and normal eyes occurred by chance.

From a clinical standpoint, high sensitivity is critical for a screening or diagnostic aid for AMD, as missing true cases could delay intervention and lead to irreversible retinal damage. While a specificity of 65% means some healthy individuals would be labeled positive and directed to follow-up testing, such false positives are less problematic if subsequent examinations (e.g., retinal imaging or comprehensive ophthalmic evaluations) can definitively rule out disease. In contrast, false negatives, particularly in early AMD, could result in lost opportunities for timely intervention. Thus, the model’s bias toward minimizing false negatives aligns with clinical best practices in diseases where early detection markedly improves outcomes.

The ROC and PR curves support the stability of this GMM-based classification system under different decision thresholds, and the high AUC-PR values in particular, are valuable in addressing the pitfalls of using ROC curves in imbalanced datasets. With AUC-PR nearing 0.97 for all AMD, the model is adept at achieving both high recall (few misses) and strong precision (few false alarms) among the positive class. This balance could prove essential in larger-scale population screening programs, where the prevalence of AMD (especially early-stage AMD) may be low, yet the cost of missing a case is high.

Overall, **these results confirm that VEP features analyzed with a Bayesian GMM can effectively distinguish both early and advanced AMD from healthy aging eyes, supported by pronounced effect sizes and near-perfect statistical power** Although specificity may require optimization or supplemental diagnostic tests, the combination of strong sensitivity and robust performance metrics underscores the potential clinical utility of this approach. If validated in broader patient populations, this methodology may help identify AMD cases at the earliest possible stage, ultimately preserving vision and improving patient outcomes.

### Comorbidities of the Patient Pool

A thorough evaluation of comorbidities across our participant categories revealed distinct ocular conditions frequently coexisting with AMD. The Early-stage dry AMD (AREDS 1) group (n=19) predominantly exhibited nonexudative AMD (91%) individuals, and presbyopia (79%). Additional frequent diagnoses included dry eye syndrome, vitreous degeneration, and nuclear sclerosis. In the Intermediate-stage dry AMD (AREDS 3) cohort (n=18), nonexudative occurred in 22 (91%), alongside presbyopia (66%). Nuclear sclerosis and cortical senile cataract also appeared notably. The Advanced AMD (AREDS 4/5) group (n=19) showed nonexudative AMD (87%), and presbyopia (58%), while 37% individuals also had exudative AMD.

Common across these categories were lens opacities, vitreous degeneration, and drusen, highlighting the multifactorial nature of age-related ocular changes. Presbyopia—an expected finding in older adults—appeared throughout. While the direct impact of presbyopia on dark adaptation remains unclear, other tests in our laboratory have shown that it can impact multi-focal VEP signals. Overall, these data underscore the complexity of AMD management: coexisting conditions can confound clinical signs, complicate therapeutic interventions, and potentially interact with disease progression. Consequently, comprehensive ocular evaluations, including assessment for cataracts, vitreoretinal pathologies, and corneal surface disorders, are crucial in effectively diagnosing and treating AMD in older populations.

### Confounders reduce specificity metrics

Additionally, while scotopic VEP effectively distinguished AMD from normal aging, its specificity against other macular diseases such as the comorbidities listed above remains to be studied. We initially tested a larger cohort of healthy participants, including younger individuals, but for the final performance metric calculations, we trimmed the dataset to include only age-matched healthy controls. Specificity was significantly higher than 0.65 with younger participants, suggesting that confounding factors in older control groups may have influenced visual stimulus perception. Further research is needed to assess the impact of these confounders on test performance.

### Future improvements

Although our study demonstrated promising accuracy, several improvements could lead to further increase in accuracy and sensitivity scores. Refraction correction using eyeglasses should be used in future studies. Technical improvements to reduce data exclusion due to signal quality are also necessary. While the group encompassed various disease stages, larger multi-site trials are necessary to confirm reproducibility. Additional improvements could include: reducing the 15-minute dark adaptation window using predictive algorithms [30]; correlating DAVEP responses with other clinical assessments, such as quantitative retinal imaging (OCT biomarkers, pigment epithelium metrics) to validate scotopic VEP as a surrogate endpoint for AMD trials; longitudinal studies to determine whether DAVEP can predict disease progression or monitor therapeutic response; and using more sophisticated machine learning classification algorithms. The clear separation of feature distributions suggests that the extracted response features are highly suitable for machine learning algorithms. However, due to the limited sample size in this study, we refrained from implementing supervised machine learning classification methods.

### Conclusions

We successfully developed a portable, objective user-friendly VEP system and an advanced Bayesian-GMM statistical analysis framework capable of identifying DAR deficits in AMD patients. This novel technology shows high potential for early AMD detection and could serve as a non-invasive, objective diagnostic tool for AMD screening in clinical and remote settings.

## Data Availability

All data produced in the present study are available upon reasonable request to the authors

## FUNDING AND ACKNOWLEDGEMENTS

This work was done thanks to our collaborators at the departments of Psychology, Physics, and Mechanical and Industrial Engineering of Northeastern University (Boston MA) and clinical staff at Advanced Eye Centers (Dartmouth, MA). This work was partially supported by grant 5R44AG057250-03 from the National Institute of Health (National Institute on Aging).

## Declaration of Generative AI and AI-assisted technologies in the writing process

During the preparation of this work the author(s) used OpenAI ChatGPT for rewording and polishing the text. AI/LLM was not used for analysis or making any figures or tables. After using this tool/service, the author(s) reviewed and edited the content as needed and take(s) full responsibility for the content of the publication.

## Notes

### Competing Interest Statement

Peter Bex, Craig Versek and Srinivas Sridhar are inventors on patents related to the NeuroVEP technology. S. Mohammad Ali Banijamali and Craig Versek were employees of NeuroFieldz Inc during the study period. Craig Versek and Srinivas Sridhar declare financial interest in NeuroFieldz Inc.

